# Lockdown Effects on Sars-CoV-2 Transmission – The evidence from Northern Jutland

**DOI:** 10.1101/2020.12.28.20248936

**Authors:** Kasper Planeta Kepp, Christian Bjørnskov

## Abstract

The exact impact of lockdowns and other NPIs on Sars-CoV-2 transmission remain a matter of debate as early models assumed 100% susceptible homogenously transmitting populations, an assumption known to overestimate counterfactual transmission, and since most real epidemiological data are subject to massive confounding variables. Here, we analyse the unique case-controlled epidemiological dataset arising from the selective lockdown of parts of Northern Denmark, but not others, as a consequence of the spread of mink-related mutations in November 2020. Our analysis shows that while infection levels decreased, they did so before lockdown was effective, and infection numbers also decreased in neighbour municipalities without mandates. Direct spill-over to neighbour municipalities or the simultaneous mass testing do not explain this. Instead, control of infection pockets possibly together with voluntary social behaviour was apparently effective before the mandate, explaining why the infection decline occurred before and in both the mandated and non-mandated areas. The data suggest that efficient infection surveillance and voluntary compliance make full lockdowns unnecessary at least in some circumstances.

## 1. Introduction

After the new Sars-CoV-2 virus arrived in early 2020, almost all Western countries implemented some form of societal lockdown, ranging from school closures to curfews.[1] Most governments reacted at the same time in similar ways, partly copying China’s approach in the Wuhan province.[2] It is a central ongoing challenge to determine which – if any – non-pharmaceutical interventions (NPIs) work best, and to what extent mandates work vs. voluntary compliance.

The similarity in the Spring response has challenged research into the effectiveness of NPIs, because of a lack of proper real control cases. This has led to a predominance of indirect modelling studies based on assumed counterfactual curves[3], [4], and empirical studies investigating points of change in country-comparisons[5].

Some epidemiological computer models suggest that NPIs such as school closures, mandated isolation, and stay-at-home orders could reduce infection rates and thus postpone deaths substantially[4]. These models require counterfactual estimates of how infection would have been without NPIs, which are highly uncertain, as discussed below.

At the same time, cross-country observational studies, although based on the empirical data, suffer from massive uncertain confounders that also change curve structure. They need to include large ensembles of countries, since there are enough few-country comparisons to favour any desired hypothesis. Erroneous conclusions are risked in particular near the tipping point of a curve partly shaped by confounders that affect natural heterogeneous transmission and susceptibility and thus curve structure [6]–[12], including population age and health status, which in some cases appear to matter more than NPIs.[13]–[16] Even when NPIs appear effective, the voluntary counterfactual to mandates (how the curve would have looked upon recommendation rather than mandate) is not easily identified.

In this paper, we approach these challenges by using evidence offered by a quasi-natural experiment in the Danish region of Northern Jutland. After the discovery of mutations of Sars-CoV- 2 in mink – a major Danish export – seven of the 11 municipalities of the region went into extreme lockdown in early November, while the four other municipalities retained the moderate restrictions of the remaining country. Incidentally, the infection numbers in the two groups are relatively comparable and counted in the hundreds, i.e., the data set has strong statistical power. This situation provides us with a unique data set of an intra-country (homogenous population) comparison with a direct case control, highly focused in both time and space (thus reducing confounding factors to the maximal extent possible).

As seen in the following, the seven municipalities did not have significantly different infection development prior to the intervention than the four municipalities that were not locked down. The infection levels decreased already at least a week before the lockdown would have any effect, and there was also a decline in case numbers in the neighbour municipalities. Test behaviour was very distinct, with the locked-down areas subject to several weeks of mass testing, but we show that this did not affect the infection dynamics, since infection curves before and after mass testing were similar in the two groups. Possible reasons behind this finding are discussed. We conclude that efficient infection surveillance prior to lockdown combined with voluntary compliance produced similar effects as mandated lockdown, although we cannot separate the relative contribution of these two effects.

## 2. Data and strategy

### 2.1. Data set and restrictions

The data consist of daily infection rates and tests performed for the Danish region of Northern Jutland (**Figure A1-A5**). We used the quasi-experiment that of 11 municipalities in the region (**Figure A1**), Aalborg, Mariagerfjord, Morsø, and Rebild continued with the national restrictions while Brønderslev, Frederikshavn, Hjørring, Jammerbugten, Læsø, Thisted, and Vesthimmerland went into extreme lockdown, announced November 5. The restrictions were effective from November 6-9 and included: Work-at-home (Nov. 6), closing of bars, restaurants, cultural and leisure activities etc. (Nov. 7), school closure and distance learning, 4^th^ to 8^th^ grade and closing of public transport (November 9). Cafés, restaurants and bars were allowed to serve takeaway. Citizens were banned from passing the municipal border with the exception of essential health workers, and all public transportation across municipal borders was shut down.

The NPIs lasted for 1-2 weeks, similar to the typical full infectivity period of Sars-CoV-2, which is thus ideal for testing short-term effects without the risk of confounders from other events.^1^ The ban on cross-municipal travel effectively also shut down a number of firms and other activities, as many employees work in other municipalities than where they live. Statistics Denmark (2020) estimated that the lockdown affected 13,600 pupils and 126,000 employees of the 280,000 inhabitants in the area.[17] The lockdown was not predicated on rising infection rates in the seven municipalities, but aimed at controlling the spread of mink-related Sars-CoV-2 mutants.

In addition to the unique focus in time and space and the equally sized test and control groups, a further advantage of the Danish dataset is the high testing regime in Denmark, which implies that most cases were identified, in fact in both groups of municipalities (See below). In addition, 2/3 of all inhabitants of the lock-down municipalities were tested during the time course [18]. The relevant case, test, and test positivity data are shown in **Figures A2, A3**, and **A4**.

### 2.2. Time considerations in identifying NPI effects

The mean incubation period of Sars-CoV-2 is 4-6 days[19]–[21] and then it takes several days of symptoms before an infection is registered. The population of actively infected needs to accommodate the impact of the NPI dynamically, making any impact of an NPI unlikely to be observed until 1-2 weeks after the effective date. Asymptomatic and pre-symptomatic cases discovered during broad testing show up earlier in the case data but they are a minority if testing is effective[22]. However, to avoid any artefact of this possibility, some effects could be considered possibly visible already November 9, i.e. at the day of the last restrictions being enforced and 3 days after the first restrictions, noting that we expect that PCR positivity cannot be measured immediately in a newly infected individual.[23] November 9 is thus an extreme early-most time point for seeing effect of the lockdown, only relevant because of the proficient testing regime in Denmark.

### 2.3. Test effects

Although accounting for test strategy is a prerequisite for analysing case numbers, with test positivity or intermediate correction schemes being reasonable in moderate testing regimes, the extreme mass testing in mid-November in the locked down municipalities (**Figure A3**) makes any measure of effect that relies on the number of tests in this period potentially spurious if cases were already well-accounted for in the standard testing regime, which we find below. If so, an incidence estimate that takes into account test numbers (e.g. test positivity) would yield excessively small corrected incidences during the days of mass testing, and then immediately higher after mass testing stopped. Such a method would be erroneous from the test data directly seen in **Figure A3**, and from the test positivity in **Figure A4**, which is very similar for the two groups of municipalities both before and after the mass testing period, but not during the two weeks of mass testing in mid-November.

**Figure A5** shows that the mass testing in mid-November was completely saturated already at 5-6,000 tests, and thus direct weighting down positives by test numbers would be erroneous in this case (but not necessarily in moderate testing regimes where test positivity is a useful indicator). The Danish data set is interesting by its focused information on test-positivity relations, notably by showing how almost complete testing affects the test-positivity relationship distinctly from “normal behaviour” as seen in the control municipalities (**Figure A5**): The two groups of municipalities do not follow the same test-positivity relationship and thus one cannot use the same test-corrected incidence (such as test positivity or a power-law correction) for them. Instead, **Figure A4** and **A5** show that the majority of cases were identified in both groups, as mass testing did not lead to more positives. For this reason, the Danish data set should be analysed with direct case numbers, not by test-corrected incidence. The rapid relaxation of test positivity after mass testing from below 0.5% back towards the level of the control group with rapid increase towards 1.5-2% in just a week (**Figure 4A**) also confirms this.

### 2.4. Modelling and statistics

In order to assess the efficacy of the lockdown implemented in Northern Jutland, we follow the infection rate per 1000 inhabitants from September 1 until November 30, 2020.^2^ Our dependent variable is the logarithm of the observed infection rate per 1000 inhabitants (plus one) while we add either the dependent variable lagged one day or the infection rate during the last seven days as our main control variables, to take into account the substantial persistence and variable time regimes in processes of community infection. We express our data in logarithms in order to reduce the influence of outlier observations, which also implies that all results can be interpreted directly as elasticities. In a set of separate tests, we additionally include the logarithm to the daily test number in each municipality.

We note that population-normalisation would give undue weight to changes in small numbers of statistical insignificance, e.g., the very small island municipality Læsø, which experienced fluctuating daily positives of only 0-1, and would count disproportionately in an effect calculation using population-adjusted incidence. At any point where the mandate could be effective, from November 9 to 30, Læsø had zero daily positives, except one day, where it had 2 (November 12), so an incidence-based effect including Læsø in this time interval alone could produce spurious effects. Instead, we consider the actual, total infections in the two groups, as is also more intuitive.

Our main independent variable is a dummy, ‘lockdown plus 7 days’, that takes the value one from seven days after the lockdown was announced, so as to account for the time it takes for infections to be detectable. We alternatively code lockdown as effective after either four or ten days, to account for the possibility that the intense broad testing may cause pre-symptomatic cases to be identified by PCR several days before symptom onset (boundary scenario used for sensitivity analysis). In separate tests, we include the spatial infection rate – i.e., the average daily infection rate in geographical neighbour municipalities – which we also alternatively lag two days, and the spatial lockdown variable, which takes care of potential cross-municipal infection chains. In the latter test, we also add an interaction term between the lockdown variable and the spatial infect rate, which allows us to assess if lockdown affected the spatial infection spill-overs. Finally, in tests in which we include the test numbers, we also add a dummy that takes the value one in the locked down municipalities from seven days before the lockdown was announced to seven days after the announcement. We do so as a placebo test because the effect of lockdown on infection rates and, in particular, on the positive ratio logically cannot occur prior to the lockdown.

The full dataset, consisting of daily infection numbers from 11 municipalities during a 91-day period, is summarised in **Table 1**. All regression analysis was performed in Stata version 16,[24] using OLS linear regression. In all tests, we added a full set of municipal fixed effects as well as week and weekday fixed effects. We thus estimate the baseline specification in 1), where Inf _t_ is the number of infected at time t, Lock is a dummy for lockdown, and D is a matrix of municipality, week and weekday fixed effects; this baseline is supplemented by additional variables, X, in the following. The municipality fixed effects control for approximately time-invariant confounders such as demographic differences, population density, local health conditions, industry structure etc. while the week fixed effects effectively control for national changes in virus policy, broad seasonal changes in weather and temperatures, as well as most confounders and other factors that affect all 11 municipalities at the same time. Finally, the weekday fixed effects reduce noise coming from, e.g., changing composition of the test population over the week.^3^ Throughout, we report standard errors clustered at the municipality level, which is necessitated by the substantial differences across the municipalities we compare in size and composition.^4^

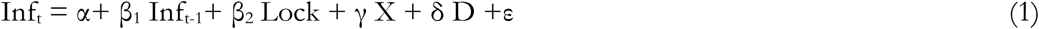

**Table 1.** Descriptive statistics.

|  | Mean | Standard deviation | Observations |
| --- | --- | --- | --- |
| Log number infected<sup>a</sup> | .079 | .098 | 1001 |
| Log number tested<sup>b</sup> | 5.601 | 1.495 | 1001 |
| Log infected last week<sup>c</sup> | .424 | .316 | 1001 |
| Lockdown plus 7 days | .133 | .339 | 1001 |
| Lockdown plus 4 days | .154 | .361 | 1001 |
| Lockdown plus 10 days | .112 | .315 | 1001 |
| Lockdown minus 14 days | .098 | .297 | 1001 |
| Spatial infected | .407 | .544 | 1001 |
| Spatial infected plus 2 days | .402 | .541 | 1001 |
| Spatial lockdown | .116 | .301 | 1001 |

In addition to the same-size test and control data sets (with / without lockdown) of neighbour municipalities, the focus in time and space and the extensive testing regime arguably make the data set unique for the purpose of evaluating relative effects of NPIs in settings of mass testing. As we show in the following, the structure of the data allows us to treat the lockdown in Northern Jutland as a quasi-natural experiment.

## 3. Results

Before turning to the empirical results, we first note that the pre-intervention infection rates did not differ between municipalities going into lockdown and those that did not. **Figure 1** provides a first impression of the development in the municipalities that would become locked down versus those that did not. Although the profiles are not identical, a set of simple balance tests provides no reason to believe that the locked down municipalities were different from those not locked down. In the former group, .15 per 1000 inhabitants tested positive per day while .14 did so in the latter group (p<.75) in the seven days before the lockdown was announced. In the following seven days, during which the lockdown most likely could not visibly affect case numbers, due to the median incubation period of Sars-CoV-2 of 5-6 days,[21] the corresponding numbers were .11 and .11 (p<.78). In the spring, the former group experienced a total of .69 positive tests per 1000 inhabitants while the latter saw .82 positive tests (p<.59). As such, we find no statistically significant differences between the two groups of municipalities prior to intervention. The strong similarity in infection rates at different timescales before the intervention strongly supports treating the lockdown as an actual quasi-natural experiment.

**Figure 1.**
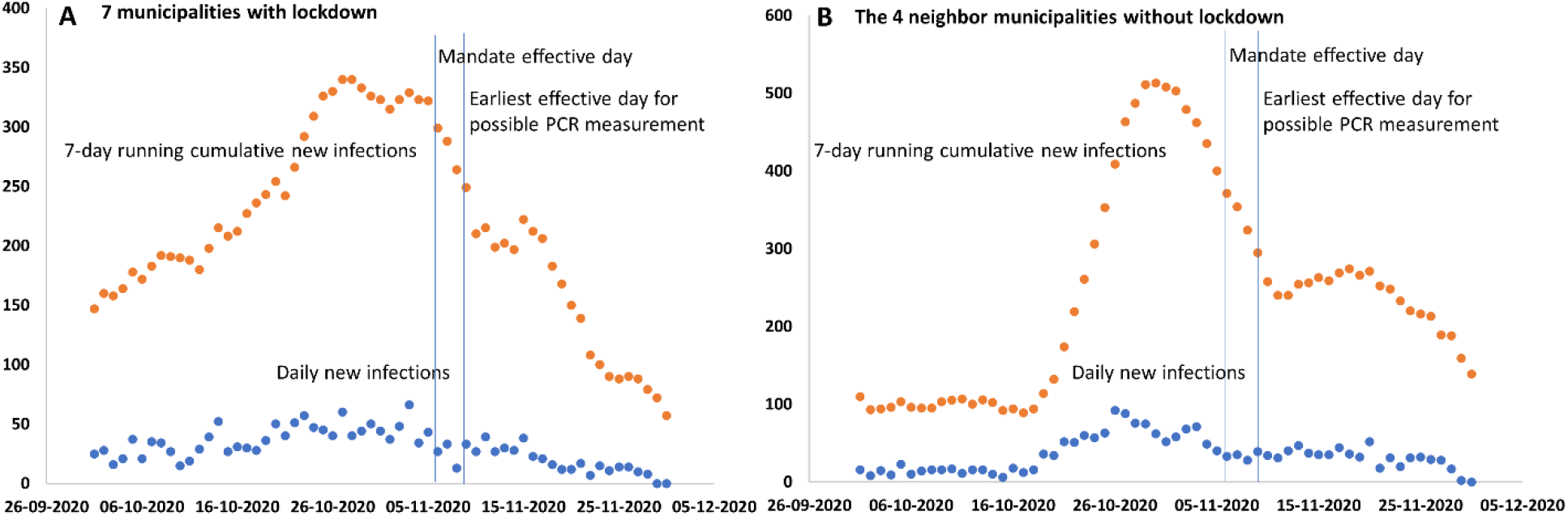
Infection development in Northern Jutland. Reported infection levels in the administrative region of Northern Jutland around the time of the November lockdown (Blue: daily new positives; orange: running weekly summed positives). (**A**) The seven municipalities with lockdown mandate. (**B**) The four municipalities without lockdown mandate. Vertical lines indicate first and last days of mandate effective (November 6 and 9). Any effect has to emerge later than this, since PCR also takes time to manifest in the population of positives.

We report the main results of our analysis in **Table 2**, where columns 1 and 2 present two different baseline specifications; column 1 uses the one-day lagged infection rate as control while column 2 uses the accumulated infection rate of the preceding week. In both cases, the lagged dependent variable is positive and significant, indicating short-term persistence. However, the specification with the weekly lagged infection rates provides a substantially better fit with the data, as indicated by the strong significance and the goodness-of-fit statistics in the lower panel. The estimate indicates that if a municipality has had ten percent higher infection numbers, relative to the common development in Northern Jutland, within the preceding seven days, it will on average see 1.4 percent more infections than otherwise. In other words, the estimate indicates that although there is short-term persistence, the size also indicates a significant regression-to-the-mean effect such that increases tend to be temporary.

**Table 2.** Main results.

|  | 1 | 2 | 3 | 4 |
| --- | --- | --- | --- | --- |
| Log infected yesterday | .295*<br>(.129) |  |  |  |
| Log infected last week |  | .148**<br>(.027) | .146**<br>(.029) | .147**<br>(.029) |
| Lockdown plus 7 days | -.055<br>(.029) | -.025<br>(.017) |  |  |
| Lockdown plus 4 days |  |  | -.033<br>(.024) |  |
| Lockdown plus 10 days |  |  |  | -.033<br>(.024) |
| Weekday FE | Yes | Yes | Yes | Yes |
| Week FE | Yes | Yes | Yes | Yes |
| Municipality FE | Yes | Yes | Yes | Yes |
| Observations | 1011 | 1011 | 1011 | 1011 |
| With R squared | .222 | .284 | .289 | .285 |
| F statistic | 50.76 | 27.60 | 27.90 | 27.80 |

Note: \*\* (\*) denote significance at $p < .01$ ( $p < .05$ ). FE refers to fixed effects. Numbers in parentheses are standard errors clustered at the municipality level. The dependent variable is the logarithm to the daily number of infected people per 1000 inhabitants. Column 1 uses the one-day lagged infection rate as control. Column 2 uses the accumulated infection rate of the preceding seven days. Column 3 has a lag of the lockdown effect of four days. Column 4 has a lag of the lockdown effect of 10 days.

Conversely, with respect to our main variable, although negative, the estimated effect of the special lockdown of Northern Jutland is far from significance at conventional levels: although the estimate indicates a 2.5 percent decrease in locked down municipalities, the 95 percent confidence interval of the estimate in column 2 is [-.063; .014]. Our baseline result thus is that the lockdown was ineffective relative to the control.

In columns 3 and 4, we instead lag the lockdown effect by either four or ten days in order to ensure that our results are not driven by the specific assumption of a one-week incubation period (if for example, the intense PCR testing captured many pre-symptomatic cases or if the lag is alternatively longer; both considered less likely boundary situations). The estimate nevertheless remains approximately unchanged and we again find no evidence of a statistically significant effect.

In **Table 3**, we study spatial spill-overs, i.e., the association between the daily infection numbers in a municipality and the corresponding numbers and lockdown status of its neighbouring municipalities, which could potentially affect the precision of our estimates and the overall infection rates. In column 1, we first add the spatial infection rate – the average infection rate in neighbouring municipalities. In column 2, we lag this spatial variable additionally two days to allow for possible delayed infection across municipal borders as a sensitivity test. None of the estimates are significant at the 95% confidence level and both are small. In column 3, we also add a spatial lockdown variable, calculated as the share of neighbouring municipalities that are locked down, in addition to the spatial infection rate. Doing so renders the spatial infection rate significant. Finally, we introduce an interaction between the spatial infection rate and the spatial lockdown variable in addition in column 4. This implies that the measured effect β_Lockdown_ of lockdown is conditional and calculated as β_Lockdown_ + β_Interaction_ * ‘spatial infection rate’; symmetrically, the effect of the spatial infection rate becomes β_Spatial infection_ + β_Interaction_ * ‘lockdown’. We mainly introduce this interaction to account for the possibility that municipal lockdowns prevent spread across municipal borders and thus break whatever infection chains that may have existed.

**Table 3.** Spatial spill-overs.

|  | 1 | 2 | 3 | 4 |
| --- | --- | --- | --- | --- |
| Log infected last week | .147**<br>(.028) | .148**<br>(.028) | .147**<br>(.028) | .124**<br>(.029) |
| Lockdown plus 7 days | -.024<br>(.017) | -.024<br>(.017) | -.025<br>(.017) | -.004<br>(.008) |
| Spatial infections | .015<br>(.008) |  | .019*<br>(.008) | .008<br>(.011) |
| Spatial infections plus 2 days |  | .006<br>(.011) |  |  |
| Spatial lockdown |  |  | .022<br>(.022) | -.022<br>(.018) |
| Spatial infections * lockdown |  |  |  | .151**<br>(.025) |
| Weekday FE | Yes | Yes | Yes | Yes |
| Week FE | Yes | Yes | Yes | Yes |
| Municipality FE | Yes | Yes | Yes | Yes |
| Observations | 1011 | 1011 | 1011 | 1011 |
| With R squared | .289 | .284 | .288 | .313 |
| F statistic | 26.02 | 25.77 | 24.58 | 26.12 |

Note: \*\* (\*) denote significance at $p < .01$ ( $p < .05$ ). FE refers to fixed effects. Numbers in parentheses are standard errors clustered at the municipality level. The dependent variable is the logarithm to the daily number of infected people per 1000 inhabitants. The conditional marginal effect of spatial infections when a municipality is surrounded by locked down neighbours is .159 (.027) and significant at $p < .01$ .

The interaction term *per se* becomes significant, which necessitates that we calculate conditional marginal effects.[25] Doing so indicates that lockdown is never significant at any observed level of spatial infection rates. Conversely, we find that the spatial infection rate becomes significant in municipalities bordering those that were locked down but remains far from significance in those that do not.^5^ In other words, the interaction results show that infection rates in municipalities under lockdown became more similar during the lockdown but did not become significantly different than those in the rest of Northern Jutland that was not locked down.

In **Table 4**, we explore another aspect of the lockdown of Northern Jutland by including the logarithm to the daily number of tests performed in each municipality. As noted in **Figure A3**, the test number increased dramatically in the locked down municipalities after the lockdown went into effect. We observe that lockdown leads to a substantial increase in tests in column 4, in which the dependent variable is the logarithm to the test number. As such, the significant effect of lockdown in column 1, where we effectively estimate the change in the positive ratio – estimating the logarithm to the infection rate, controlling for the logarithm to the test number is equivalent to estimating the positive ratio (infections / tests) – is a consequence of controlling for the test strategy. The full effect of lockdown can be calculated as -.043 + .039 * .470 ≈ -.024, i.e., essentially the estimate in **Table 2**.

**Table 4.** Results including test numbers.

|  | 1 | 2 | 3 | 4 |
| --- | --- | --- | --- | --- |
| Log infected last week | .114***<br>(.029) | .112***<br>(.027) | .108***<br>(.025) | .875***<br>(.189) |
| Log number tested | .039***<br>(.009) | .042***<br>(.008) | .048***<br>(.007) |  |
| Lockdown plus 7 days | -.043***<br>(.016) | .002<br>(.035) | .011<br>(.041) | .470***<br>(.139) |
| Lockdown plus 7 * tested |  | -.008*<br>(.004) | -.012**<br>(.005) |  |
| Lockdown minus 14 days |  |  | .041**<br>(.016) |  |
| Lockdown minus 14 * tested |  |  | -.013***<br>(.002) |  |
| Weekday FE | Yes | Yes | Yes | Yes |
| Week FE | Yes | Yes | Yes | Yes |
| Municipality FE | Yes | Yes | Yes | Yes |
| Observations | 1011 | 1011 | 1011 | 1011 |
| With R squared | .347 | .349 | .358 | .312 |
| F statistic | 34.55 | 32.62 |  | 31.54 |

Note: \*\* (\*) denote significance at $p < .01$ ( $p < .05$ ). FE refers to fixed effects. Numbers in parentheses are standard errors clustered at the municipality level. The dependent variable in columns 1-3 is the logarithm to the daily number of infected people per 1000 inhabitants while the dependent variable in column 4 is the daily number of tested inhabitants.

The data for test numbers allow us to explore how the test strategy affects the observed positive ratio. We do so in column 2, where we interact the lockdown with the test number. This shows how the observed effect of the lockdown is increasing in the number of test subjects, which should not be the case if the composition of the test population was held constant. Furthermore, interacting a variable that captures the 14 days prior to the time a lockdown could have had influence on the infection rate shows the exact same heterogeneous association with the infection rate as the period after the lockdown took effect. We consider this an intuitive placebo test of the lockdown, which it fails. **Figure A4-A5** provide a perhaps more intuitive picture showing that testing is saturated at levels much below the mass testing and thus, that the groups are insignificantly different under most comparisons except those that are distorted by the mass testing in mid November, and the two groups do not follow the same test-positivity relations for this reason, and thus cannot be studied by a single test-corrected incidence together.

The basic non-result of lockdown efficacy is therefore a constant feature across all three tables and it is also robust to a series of additional tests. The estimate of lockdown varies in a full jackknife test – a set of 11 tests in which we exclude one municipality at a time – between -.034 and -.008. This test for example takes care of the potentially problematic influence of either the very small municipality of Læsø or the large municipality of Aalborg, although we find the only visible change in the estimate when excluding the municipality of Morsø. In that case, the estimated effect of lockdown drops to - .008. All of these estimates are far from significance at conventional levels. The estimate also remains insignificant and of the same size if we restrict the sample to only including observations from the later autumn (October to November), if we exclude all days without any positive tests in a given municipality, if we express infection rates in absolute numbers instead of logarithms, or if we exclude all days with either no or particularly many positive tests.^6^ The lack of efficacy is therefore not the result of noisy single observations, outlier municipalities or misleading temporal comparisons.

## 4. Discussion

Almost all Western countries used societal lockdown as a primary policy instrument in 2020. While surely having some effect on viral transmission, the specific gain of a full lockdown vs. voluntary behavioural changes and other simultaneously enforced milder NPIs remains a matter of debate due to lack of actual empirical control cases for the same populations. Modelling studies, despite the important insights they give into infection transmission and control, suffer from uncertain counterfactual curves, and country-comparisons, although analysing epidemiological data, suffer from major confounder effects.

In this study, we take advantage of a quasi-natural experiment that occurred in the Danish region of Northern Jutland in the late Autumn of 2020. Owing to a potential mutation of Sars-CoV- 2 in mink, seven of the 11 municipalities in the region went into extreme lockdown by November 6, which included a travel ban across municipal borders, closing schools, the hospitality sector and other venues. The municipalities going into lockdown did not exhibit higher infection rates prior to the lockdown and can therefore be compared to those that remained relatively open. We first and foremost alert the community to the existence of this dataset and invite other views on it with open eyes, as it is to our knowledge the most time- and space-focused empirical dataset available with sufficient statistical power, adequate and homogeneous control group, nearly complete testing, and with the smallest possible confounder pollution imaginable in a real setting.

In our own analysis of this dataset, which we invite others to investigate, we did not find evidence of any effects of the lockdown on the development of infection rates across Northern Jutland, relative to the control. In other words, we find that an extreme version of societal lockdown had no effect on virus development: Although infections fell over time, they did so before the mandate was implemented (and even before it was announced), and the decline experienced in Aalborg, the main driver of the case control group without mandate, was comparable.

Test-corrected incidences (test positivity or intermediate methods such as power law fits), as sometimes appropriate at moderate testing levels, is shown to be inadequate in this case because 1) testing was saturated and almost all cases were identified much below mass testing levels and 2) the two groups did not follow the same test-positivity behaviour during the 2 weeks of mass testing, but do so before and after, and 3) test positivity rapidly converged back to the control group levels once mass testing stopped in the lockdown municipalities (**Figure A3-A5**). As a final comment, we also looked at the infection levels during December (**Figure A2**). From their lows, both groups of municipalities experienced a rapid 5-fold increase in PCR positives that confirms our general conclusion that the overall infection dynamics in the two groups of municipalities have not been greatly affected, neither short-term, nor longer-term.

Our results question the efficacy of a full lockdown in the complex interplay with voluntary behaviour, efficient test-based infection control, and already implemented milder NPIs. We note that infection levels did decrease substantially and already before lockdown and in both groups, so something was apparently effective in Denmark. We suggest the following explanations:

1. Denmark has a very high level of testing per capita. As the data suggest that almost all real cases were identified at lower test levels, i.e., testing was “saturated” (**Figure A5 A**), our data are not sensitive to changes in test intensity as other case-based studies may be, except if erroneously using the direct mass testing test numbers to produce a test-corrected incidence, which we argue is wrong in this case (but reasonable under moderate testing regimes where the two groups have similar test-positivity relationships). Infection levels declined prior to the mandate, indicating that the infection pockets were efficiently identified and isolated before the mandate. Thus, our study may suggest that when infection levels are low enough, testing, as in the Danish context, is more important than lockdown in controlling infection. Although clearly part of the answer, we do not believe this can be the full answer since the infection tracking by PCR notoriously tails real infections by days or weeks so other effect are probably at play as well.
2. Second, voluntary compliance, trust, and general information quality is arguably very high in Denmark, which could induce a pre-mandate awareness and voluntary compliance that could have helped the specific outbreak. The fact that the municipalities without mandate declined to the same extent as the mandated municipalities, and that the decline began before the mandate was announced, indicate such type of effect at play, although it cannot be separated completely from the effect of efficient contact tracing mentioned above.
3. Third, in a more general context, it may be that lockdown effects have been overestimated. Early computer modelling indicated that mandated lockdown would be highly effective and prevent hundreds of thousands of deaths.[4] However, the homogenous models used to produce counterfactual curves have been criticized in many papers, since heterogeneity in susceptibility, activity, infectivity, and compliance all tend to flatten curves by themselves relative to counterfactual homogeneous populations [6]–[8], [10], [12]. To the extent that test-driven infection control or voluntary compliance do not fully explain our data, our results are consistent with this criticism and with country-comparison studies showing smaller direct effects of full lockdown itself[16].

Our results are broadly in line with previous studies showing that lockdown and to some extent some other strict NPIs matter less in context of efficient but milder NPIs and confounding population health variables[13]–[16], [26], [27] but this is to our knowledge the first time it is shown with a control-group dataset in a real population. According to the analysis by Chaudhry et al., full lockdowns and a high rate of COVID-19 testing did not relate to reductions in critical cases or mortality.[13]

The Danish selective lockdown illustrates an important point discussed by Haug et al.[24]: That complete lockdown is not necessarily the most effective measure in the context of other, less costly NPIs, in particular if considering the effects of infection tracing and voluntary compliance, as is probably quite good in Denmark. Our results are also resonate with those by Brauner et al.[3] who found that limits on large gatherings (100+) and closing schools and universities alone brings down R_t_ considerably, without any need of business closures or very intrusive measures such as smaller gatherings and stay at home orders. Our case-controlled single-population study also confirms the indications of Goolsbee and Syverson that voluntary behavior can overrule enforced restrictions.[28]

Our results (and most of the above epidemiological retrospective studies, in fact) are at odds with some modelling studies perhaps best illustrated by the study of Flaxman et al.[4] In addition to other points raised[29], these models lack the heterogeneity of real populations (the fact that most transmission occurs in a minority of the population, “superspreading”, etc.), which substantially reduces transmission and (transient) herd immunity thresholds[6]–[12]. Thus homogeneous models substantially overestimate the size of counterfactual curves and therefore, effects of NPIs.[7], [8] This heterogeneity both manifests as biological susceptibility and infectivity (differences in human immune responses, symptoms, and virus load) and social activity (connectivity in social networks, where superspreaders, once infected, substantially dampen transmission). These real features of populations flatten curves (although not necessarily longer-term herd immunity thresholds) before any NPI[35]. Finally, the assumption that IFR values based on seroprevalence count all infection (i.e. the assumption that all infected have antibodies) is probably wrong[36] and thus using sero-based IFR as a proxy for mortality could also overestimate NPI impact, beyond neglect of heterogeneity. Neglect of both biological and social heterogeneity as done by Flaxman et al. makes the counterfactual infections and deaths too large for both reasons[11].

A new multi-strategy study recently described well the diversity of impacts of NPIs relative to each other, of course with the limitations of confounders between country comparisons that prevail in all such analyses[3]. The additional impact of a full lockdown on top of already mandated NPIs in Denmark at the time and voluntary social distancing and sanitation behaviour of informed citizens is arguably dependent on the infection levels and testing capacity so there is no simple answer to these questions, and we need to study the evidence further in the light of the weaknesses and strength of each approach.

## 5. Conclusions

We analysed a new unique data set arising from the selective mandated lockdown of 7 municipalities of the administrative region of Northern Jutland in Denmark, but not 4 others, as a consequence of spread of mink-related mutations in November 2020. The data set has statistical power (significant change in infection during the time of study; similar sized test and control groups both with hundreds of infection cases), focus in time and space (November 2020; specific Northernmost region of Denmark considered), and highly efficient testing with data showing that testing was saturated, i.e. most cases found at the test levels of the municipalities, with little additional benefit of mass testing. We alert the world to the existence of this data set and invite additional analysis of it.

Our analysis shows that while infection levels decreased, they did so before the mandate was announced, and the restrictions also had limited and statistically insignificant effects relative to neighbour municipalities without mandates, where infection levels also decreased markedly in Aalborg, the most important municipality not subject to lockdown. A direct spill-over effect to neighbour municipalities was not seen.

As possible explanations for the results, we propose that efficient tracing of infection pockets was apparently effective and complete at the time of the mandate, and this brought down infection together with alert social behaviour that was insignificantly different between the mandated and non-mandated areas. As a partial additional explanation, the impact of full lockdowns may have been overestimated by use of homogenous population counterfactuals. Our study suggests that efficient infection tracing and voluntary behaviour is more important than actual mandates in controlling infection, at least in the Danish context studied.

## Data Availability

All data and statistical codes (the do-file) are available from the authors upon request. The case and test numbers of the 11 municipalities can be found at the Danish Statens Serum Institut (SSI) https://covid19.ssi.dk/overvagningsdata/download-fil-med-overvaagningdata

https://files.ssi.dk/covid19/overvagning/data/data-epidemiologiske-rapport-24122020-b9x0

---

**Figure A1.**
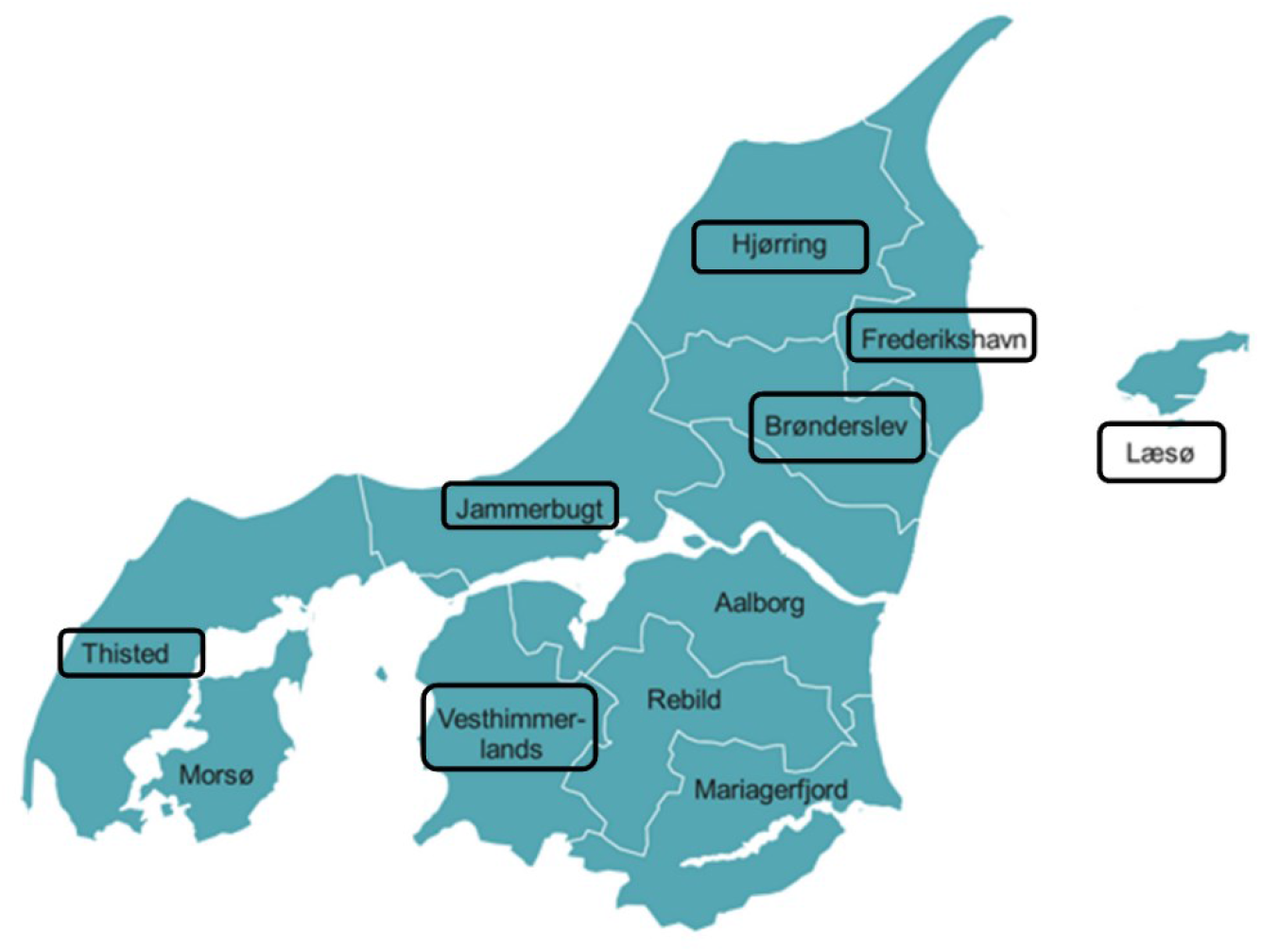
Map of Region Nordjylland of Denmark. (encircled municipalities were under lockdown.) (https://www.kl.dk/politik/kkr/kkr-nordjylland/)

**Figure A2.**
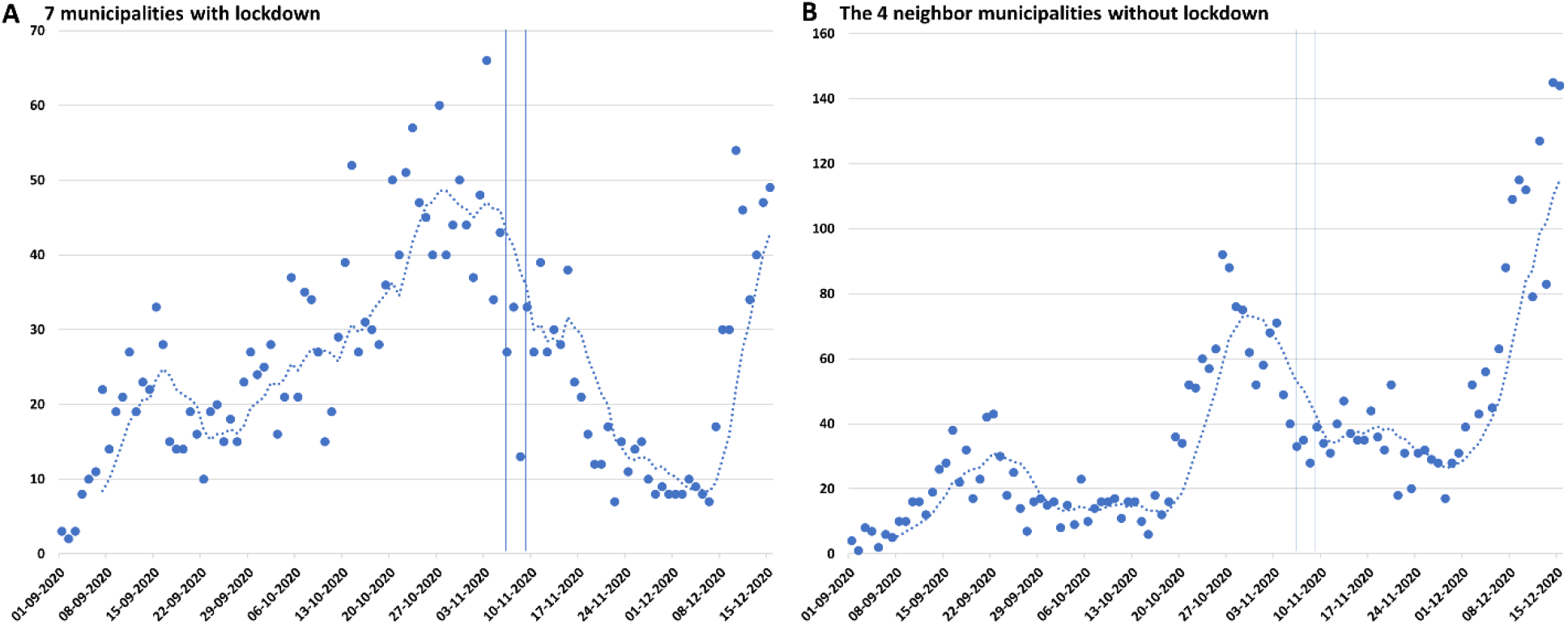
Increase in infection in December. (**A**) Municipalities with lockdown. (**B**) Municipalities without lockdown. The dashed lines are 7-day running averages. Vertical lines indicate day of mandate effective (November 6) and first day where PCR positives can possibly be registered (three days). This shortest possible interval requires almost perfect test intensity. The increase in infection in December is very similar percentwise in both groups (approximate 5-fold increase), confirming similarity also post-NPI within the noise.

**Figure A3.**
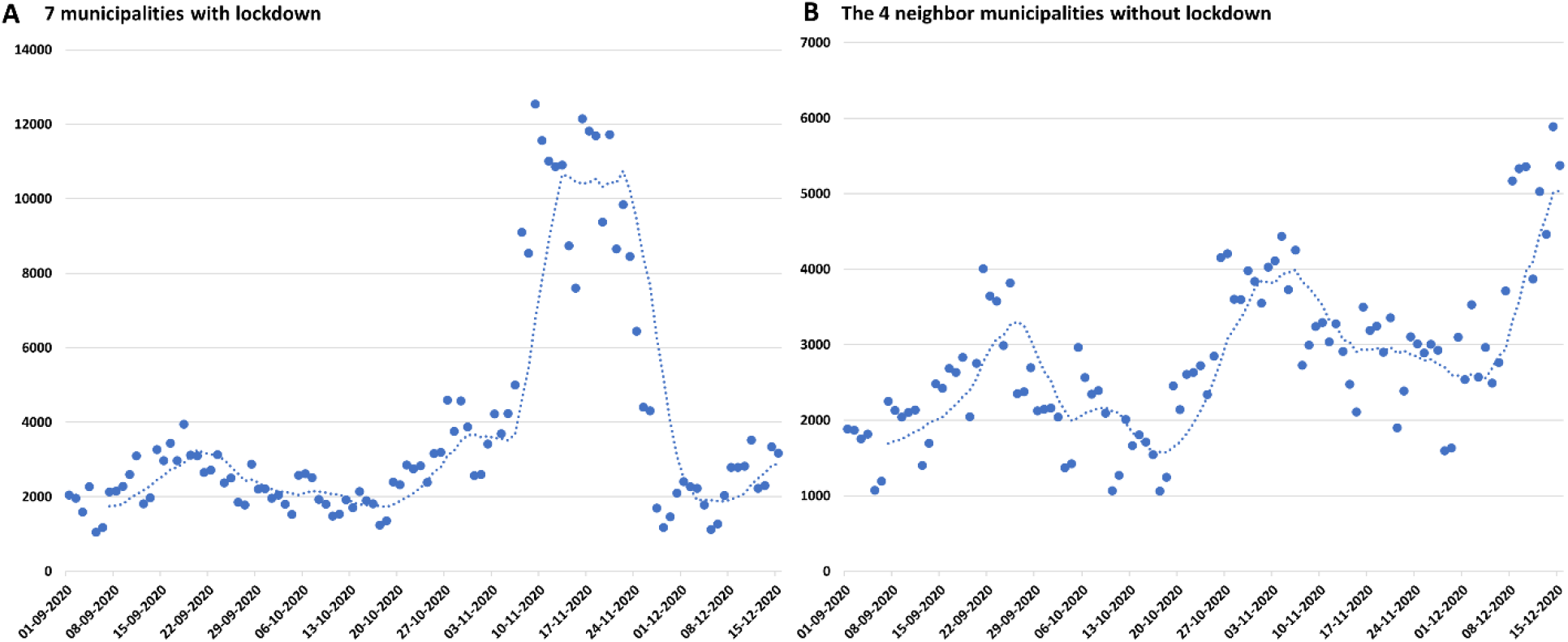
Tests performed. (**A**) Municipalities with lockdown. (**B**) Municipalities without lockdown. The dashed lines are 7-day running averages.

**Figure A4.**
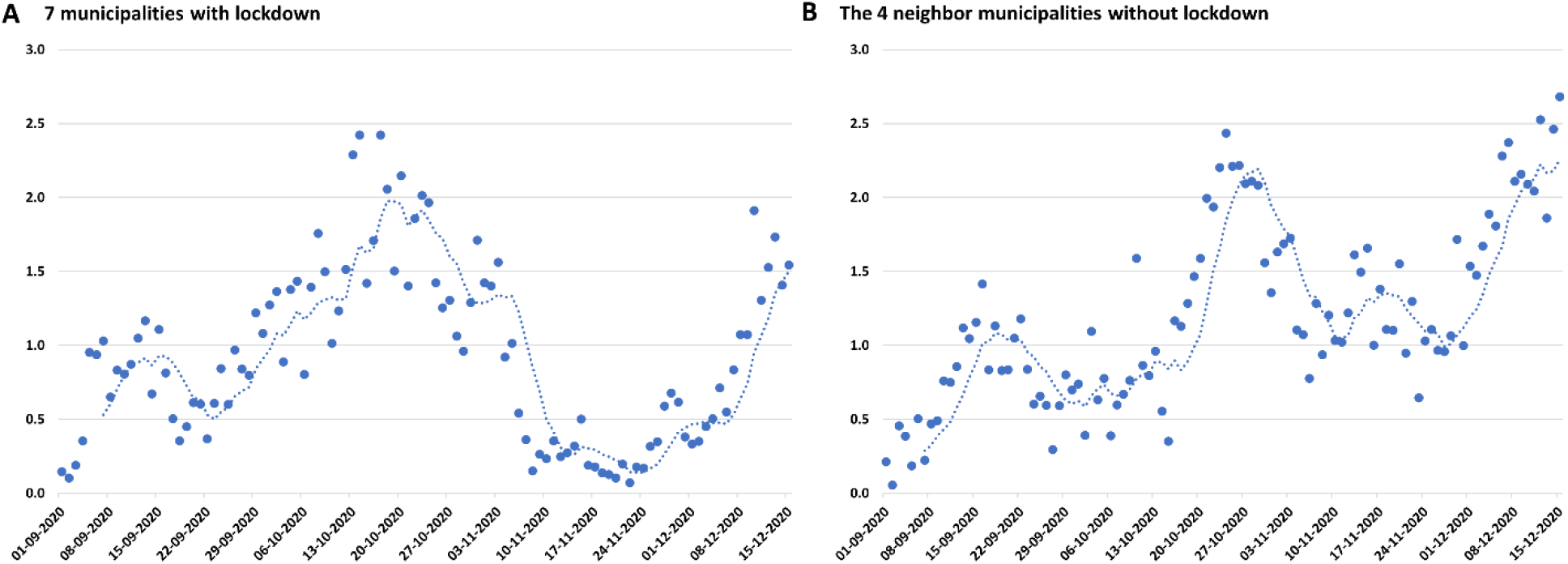
test positivity (positives / tests). (**A**) Municipalities with lockdown. (**B**) Municipalities without lockdown. The dashed lines are 7-day running averages.

**Figure A5.**
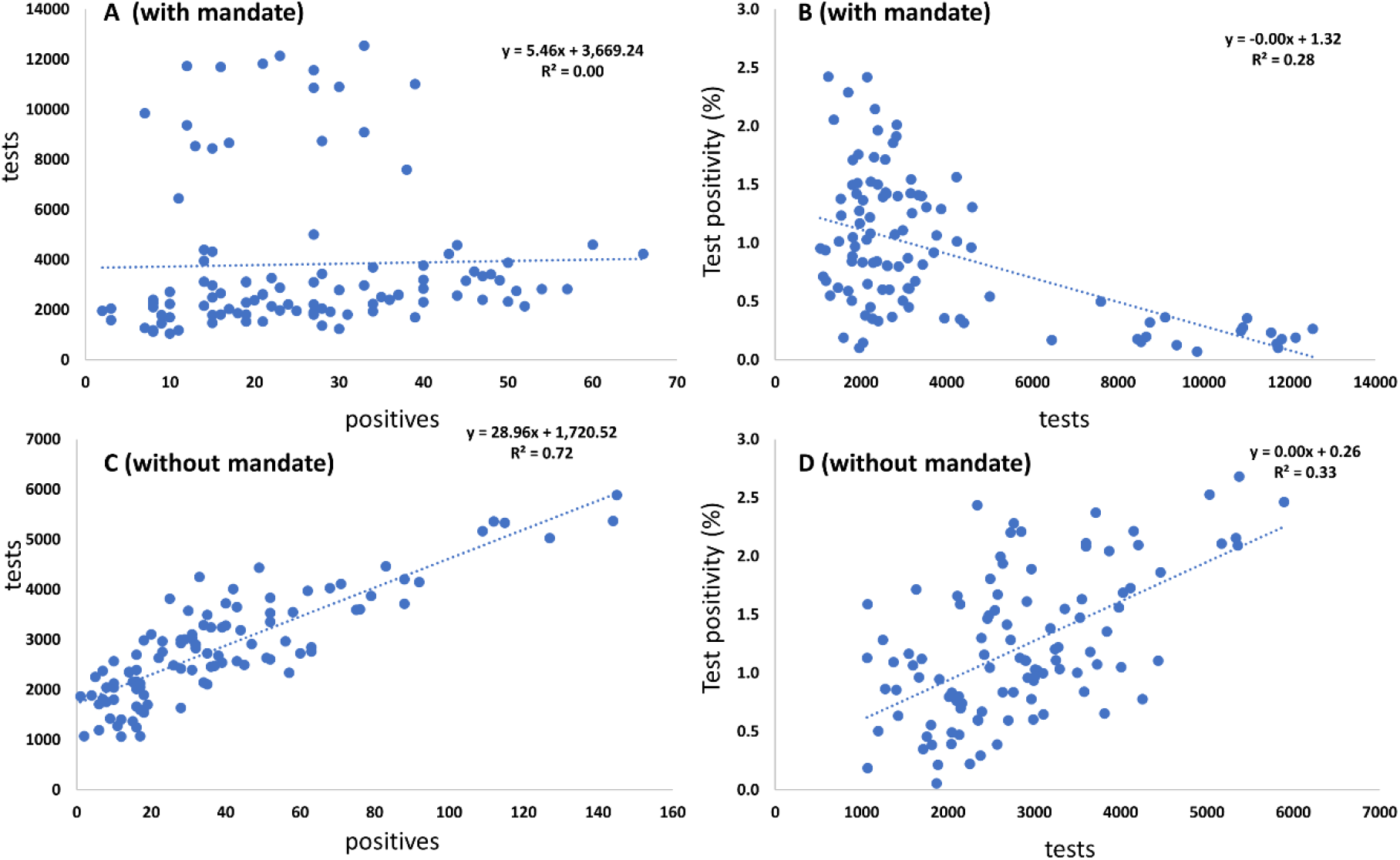
Relations between test number and positives, and test positivity (positives / tests). (**A**) Tests vs. positives for municipalities with lockdown. (**B**) Test positivity vs. tests for municipalities with lockdown. (**C**) Tests vs. positives for municipalities without lockdown. (**D**) Test positivity vs. tests for municipalities without lockdown.

## Footnotes

1 As of November 16, the health authorities allowed public transport between the locked down municipalities and allowed schools to reopen.

2 The population size of each municipality (2020): Aalborg, 217,075; Brønderslev, 35,304; Frederikshavn, 59,654; Hjørring, 64,483; Jammerbugten, 38,324; Læsø, 1,775; Mariagerfjord, 41,800; Morsø, 20,247; Rebild, 30,113; Thisted, 43,423; and Vesthimmerland, 36,727.

3 Although we do not report these results, we consistently find indications of lower measured infection rates on Fridays and Saturdays.

4 All data and statistical codes (the do-file) are available from the authors upon request.

5 This specific interaction result may indicate that infection rates in municipalities under lockdown became more similar during the lockdown but did not become significantly different than those in the rest of Northern Jutland further away from the locked down area. This finding could be interpreted as an effect of the NPI on compliance towards common (homogeneous) behaviour. Although the idea that NPIs homogenizes behaviour is intuitively appealing, an additional test in which we calculated a spatial lockdown variable *as if* neighbouring municipalities had been locked down two weeks earlier than the actual date indicates a slightly weaker but qualitatively similar effect *prior* to the lockdown, i.e. these municipalities were relatively similar before lockdown.

6 The last robustness test is a standard test in the social sciences to ascertain that extreme values of the dependent variable do not drive the results. Excluding all zero observations as well as the five or ten percent largest values yields an estimated lockdown effect around -.005. Expressing infection rates in absolute shares per 1000 inhabitants instead of logarithms results in clearly less precisely identified effects, but yields qualitatively the same implications.

